# Saliva-based COVID-19 Rapid Antigen Test: a practical and accurate alternative mass screening method

**DOI:** 10.1101/2022.10.24.22278691

**Authors:** Idrissa Diawara, Samir Ahid, Leïla Jeddane, Soyoun Kim, Chakib Nejjari

## Abstract

As SARS-CoV-2 and its variants continue to spread, a reliable and convenient alternative to nasopharyngeal swabbing and RT-PCR testing is needed. To test the usability and performance of saliva sample collection, saliva, nasal and nasopharyngeal swab specimens were collected from a total of 338 individuals consisting of confirmed COVID-19 patients and healthy subjects. To evaluate the diagnostic accuracy of self-collected and performed SARS-CoV-2 rapid antigen test on saliva and nasal swabs specimens, we compared its performance to nasopharyngeal swab specimen RT-PCR as a comparator test. In saliva specimens, the positive percent agreement was 90.14%, and the negative percent agreement was 99.61%, while in nasal swab specimens, the positive percent agreement was 91.55%, and the negative percent agreement was 100%, both meeting the sensitivity and specificity criteria required by the FDA. Therefore, when considering both the reliability and convenience of testing, we found saliva testing to be the better method for large-scale and frequent self-testing.

## BACKGROUND

COVID-19 continues to strain public health systems and alter lives around the world, as SARS-CoV-2 and its variants are rapidly spreading around the world [1,2]. Therefore, inexpensive, scalable, and sustainable strategies for accurate and rapid diagnosis of SARS-CoV-2 are in dire need, in order to effectively manage the spread of the pandemic [3,4].

Nasopharyngeal swab (NPSs) and RT-PCR based diagnosis has been globally accepted to be the gold-standard because of its role in detection of other upper respiratory tract pathogens, as well as the high sensitivity and specificity [5]. To date, hundreds of millions of individuals worldwide have been tested for SARS-CoV-2 using PCR methods [6]. However, the PCR method entails a long waiting period until detection results become available, as well as the services of skilled professionals and expensive equipment [7]. In addition, as the testing guideline first relied solely on RT-PCR testing centers, they faced intense pressure around the world, and demand for swabs and personal protective equipment (PPE) required by the healthcare professionals for sample collection drove a cascading collapse of supply chains and caused shortages of these required items [8,9].

To alleviate the reliance on RT-PCR testing, rapid self-detection methods have been explored as alternatives. Antigen detection using lateral flow-based rapid diagnostic tests (Ag-RDT) has been widely used for on-site mass diagnosis for early detection of SARS-CoV-2 [10]. However, the performance of several commercial RDTs was found to be highly variable, and without additional methods to enhance their sensitivity, the tests are limited in their use for diagnosis [11].

The use of saliva for SARS-CoV-2 detection was soon explored and promptly found to be potentially a more affordable and less invasive self-testing method without the need of skilled personnel or supply demands of swabs and PPE [12-14]. Despite the many advantages of using saliva, it has been at first controversial surrounding its sensitivity. The problem is purported to be that collection and processing methods of saliva were not standardized compared to NSP, leading to conflicting results. In addition, early studies used inpatient saliva, which is usually more viscous, making the results difficult to translate to the general population [15].

However, subsequent studies of saliva testing have been successful. Several studies have reported that saliva can serve as a transient medium for the transmission of SARS-CoV-2, which is broadly enriched on the epithelial cells lining the oral cavity and oral mucosae [16–18]. More recently, the U.S. Food and Drug Administration and the Centers for Disease Control and Prevention have updated their guidelines to include saliva-based COVID-19 testing, and saliva testing has gained traction in situations such as in educational institutions and [19,20] with other countries such as Germany and Japan following suit. The use of saliva is ideal for frequent, repeat testing and is well suited for detecting SARS-CoV-2 during the prodromal phase to curb further transmission [21].

Moreover, with the advent of B.1.1.529 (omicron) variant, first identified in November 2021 and quickly the dominant variant [22], saliva testing shows immense promise, as the omicron variant is theorized to replicate better in the mouth and the throat. Previous study evaluated the relative performance of saliva and mid-turbinate swabs for RT-PCR testing for the delta and the omicron variants and found that saliva samples outperform in detecting the omicron variant [23].

The purpose of this study was to evaluate the diagnostic specificity and sensitivity of the PCL COVID19 Ag Gold kit for self-testing by lay person use. The results of the study show that the sensitivity (90.14% and 91.55% for saliva and nasal swab) and specificity (99.61% and 100% for saliva and nasal swab) met the acceptance criteria required by FDA. This study is also the first to include children of age 2 to 18, showing the convenience and usability of the self-test saliva kit in comparison to the more invasive NSP sample collection. In conclusion, we show that the PCL COVID19 Ag Gold kit is a more convenient and noninvasive self-testing method with comparable sensitivity and specificity to the RT-PCR method, making it the inexpensive, scalable, and sustainable solution needed at this stage of the COVID-19 pandemic.

## RESULTS

**TABLE 1.** Baseline characteristics of the patient enrolled in the clinical study.

| Specimen type | Saliva | Nasal |
| --- | --- | --- |
| Total number | 329 | 328 |
| Age (years) | N |  |
| 2~13 | 28 | 30 |
| 14~24 | 76 | 75 |
| 25~64 | 171 | 170 |
| ≥65 | 54 | 53 |
| Sex | N (%) |  |
| Male | 45.9 | 45.1 |
| Female | 54.1 | 54.9 |
| Onset days of COVID-19 | N |  |
| 0~3 | 103 | 103 |
| 4~7 | 58 | 57 |
| Asymptomatic | 168 | 168 |

**TABLE 2.** Evaluation of the diagnostic performance of PCL COVID19 Ag Gold.

| Specimen type | Sensitivity (%)<br>(95% CI) | Specificity (%)<br>(95% CI) |
| --- | --- | --- |
| Saliva | 90.14%<br>(80.74%≤95% CI≤95.94%) | 99.61%<br>(97.86%≤95% CI≤99.99%) |
| Nasal | 91.55%<br>(82.76%≤95% CI≤96.07%) | 100 %<br>(98.52%≤95% CI≤100%) |

The clinical performance of the PCL COVID19 Ag Gold was evaluated in comparison with the RT-PCR test for saliva and nasal swab specimens. In saliva specimens, the positive percent agreement is 90.14% (80.74%≤95% CI≤95.94%) and the negative percent agreement is 99.61% (97.86%≤95% CI≤99.99%). In nasal swab specimens, the positive percent agreement is 91.55% (82.76%≤95% CI≤96.07%) and the negative percent agreement is 100 % (98.52%≤95% CI≤100%).

In total 47 symptomatic patients with COVID-19 positive results, Ct value in NP swab samples correlated with symptom onset days (r=0.45, p<0.01). In Figure 2A, both positive rate of rapid antigen test for saliva and nasal samples showed 100% detection rate, when RT-PCR Ct value was below 20. The Ct value was in the range of 20 to 30, the positive detection rate of rapid antigen test was 97.67% and 97.83% for using saliva and nasal samples. However, the positive detection rate decreased to around 60% (62.5% for saliva and 58.33% for nasal) when Ct value was higher than 30. In Figure 2B, within 7 days of symptom onset, higher than 90% sensitivity (91.67%) of rapid kit was detected for both saliva and nasal samples. For symptomatic patients (n=47), both positive rates using saliva and nasal samples showed 95.74% (45/47) sensitivity. Even for asymptomatic patients (n=24) confirmed with COVID-19 positive, around 80% sensitivity was detected using saliva or nasal samples (as shown in Figure 2C).

**Figure 1.**
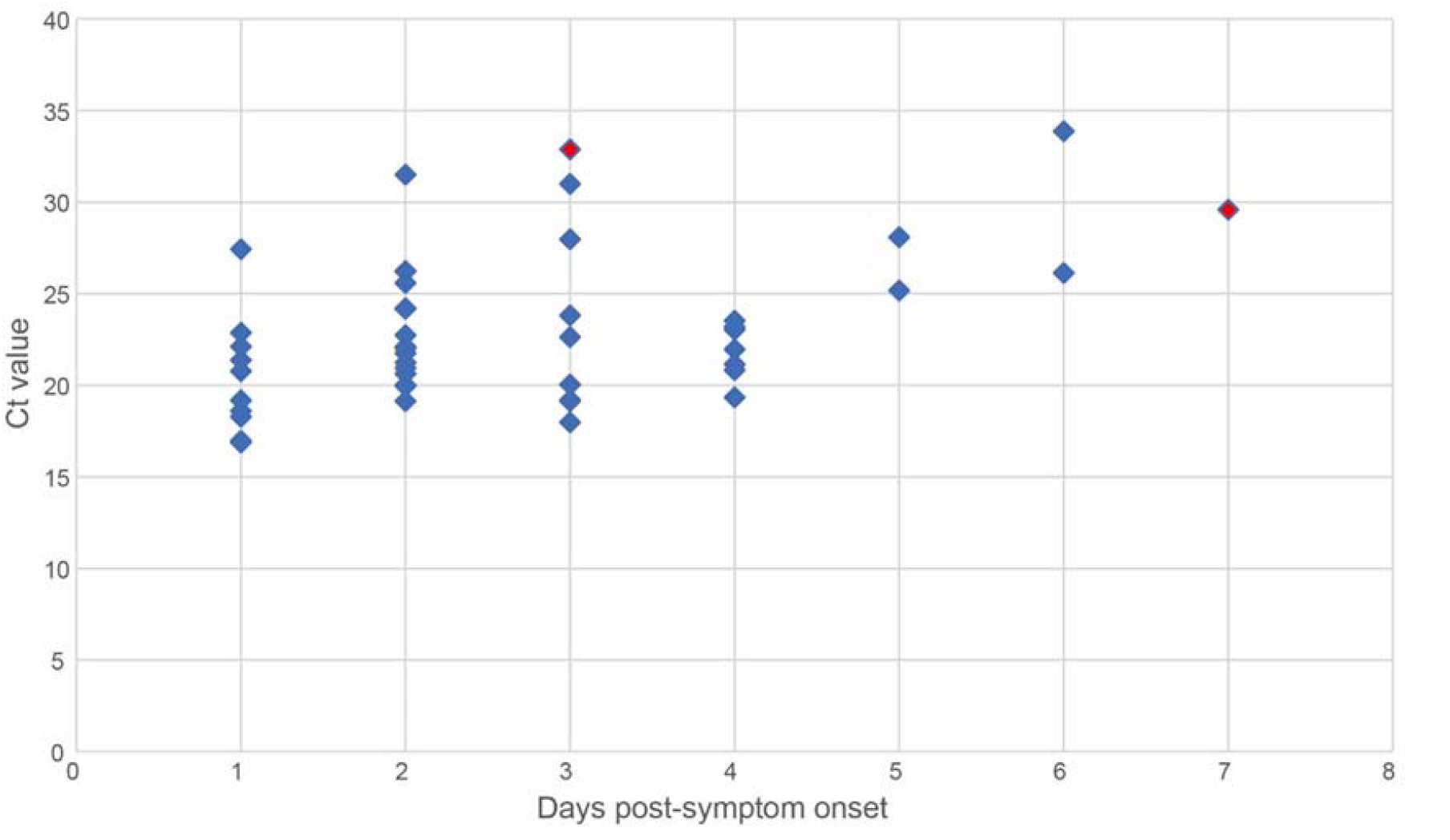
**Detection of SARS-CoV-2 in nasopharyngeal (NP) swab specimens using RT-PCR. The samples were from symptomatic patients who were confirmed with COVID-19 (n=47). Dot colors represent false-negative (red) and true-positive (blue) results by rapid antigen detection test. The correlation between Ct values of NP and the days post-symptom onset (n=47, r=0.45, p<0.01)**.

**Figure 2.**
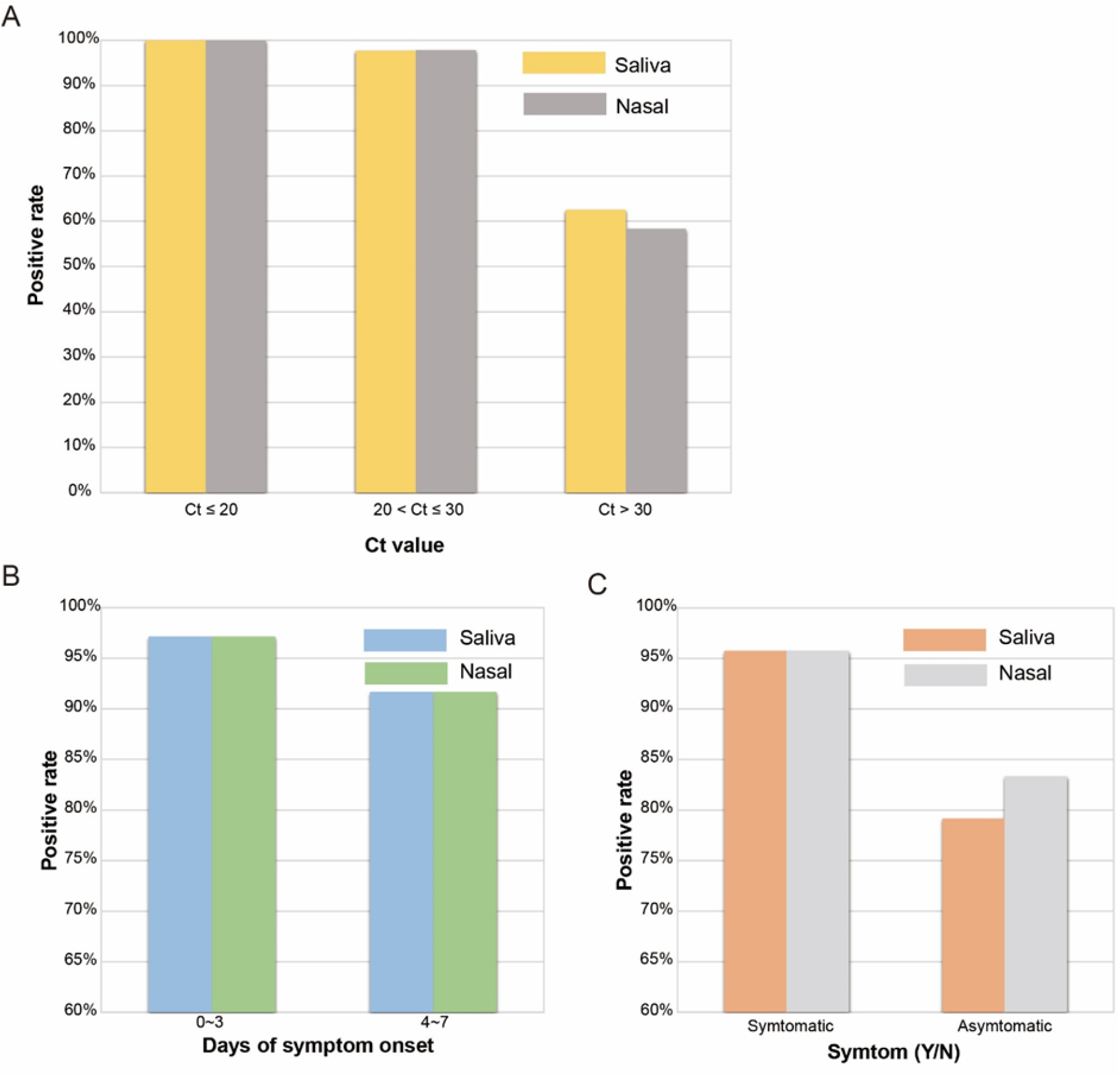
**Positive detection rate of SARS-CoV-2 in PCL COVID19 Ag Gold (COV04S): Saliva (n=71) and nasal (n=71) specimens were collected from 77 patients confirmed with COVID-19 patients. Patients were grouped according to (A) RT-PCR Ct values: Ct≤20, 20<Ct≤30, Ct>30, (B) days of symptom onset: 0∼3, 4∼7, and (C) with or without symptoms**

The median RT-PCR Ct values were 22.80 and 22.86 for the saliva positive rapid antigen test and nasal positive rapid antigen test respectively. Compared to the median RT-PCR Ct values of saliva (Ct= 32.17) and nasal (Ct= 32.53) negative rapid antigen test result, the average Ct values were significant lower in both saliva and nasal positive rapid antigen tests (P<0.001) as shown in Figure 3. In the clinical trial period, the positive rate of RT-PCR was 21.6% (71/329) and the positive rate of rapid antigen was 19.5% and 19.8% (64/329, saliva samples, 65/328 nasal samples). In Figure 4, the PCL COVID19 Ag Gold kit showed sensitivity of 100% (with Ct_≤_25) and 92.86% (with 25<Ct_≤_30) respectively. For low positive samples, both saliva and nasal rapid antigen results showed less than 60% sensitivity.

**Figure 3.**
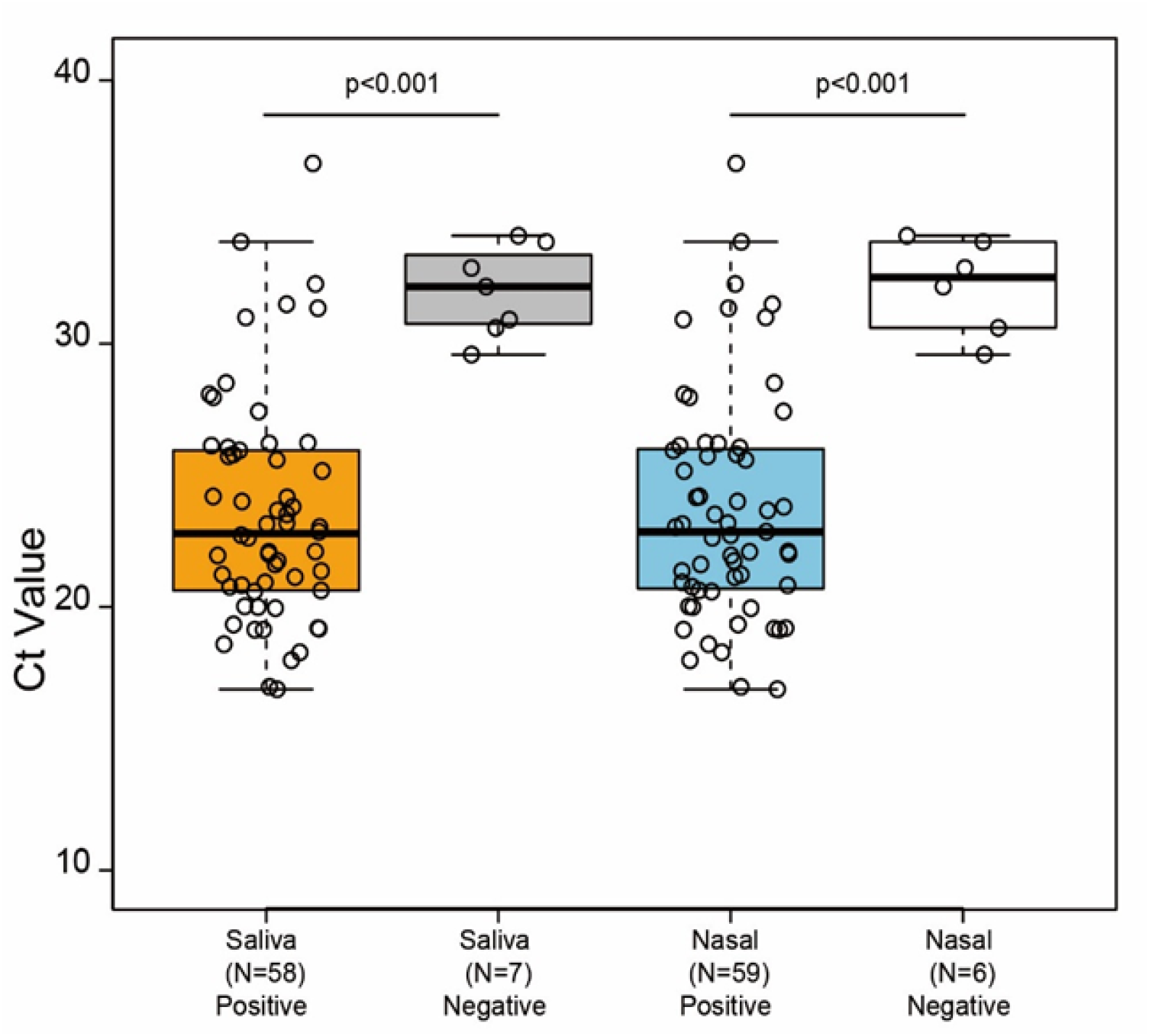
**Ct values of positive/negative results by rapid antigen test using saliva and nasal samples (n=65). In both saliva and nasal samples applied to rapid antigen test, the average Ct values were significantly lower in all positive rapid tests compared with negative rapid tests (P0.001). In the box and whisker plot, the central box represents the interquartile range, with the central line being the median, the whiskers represent the range**.

**Figure 4.**
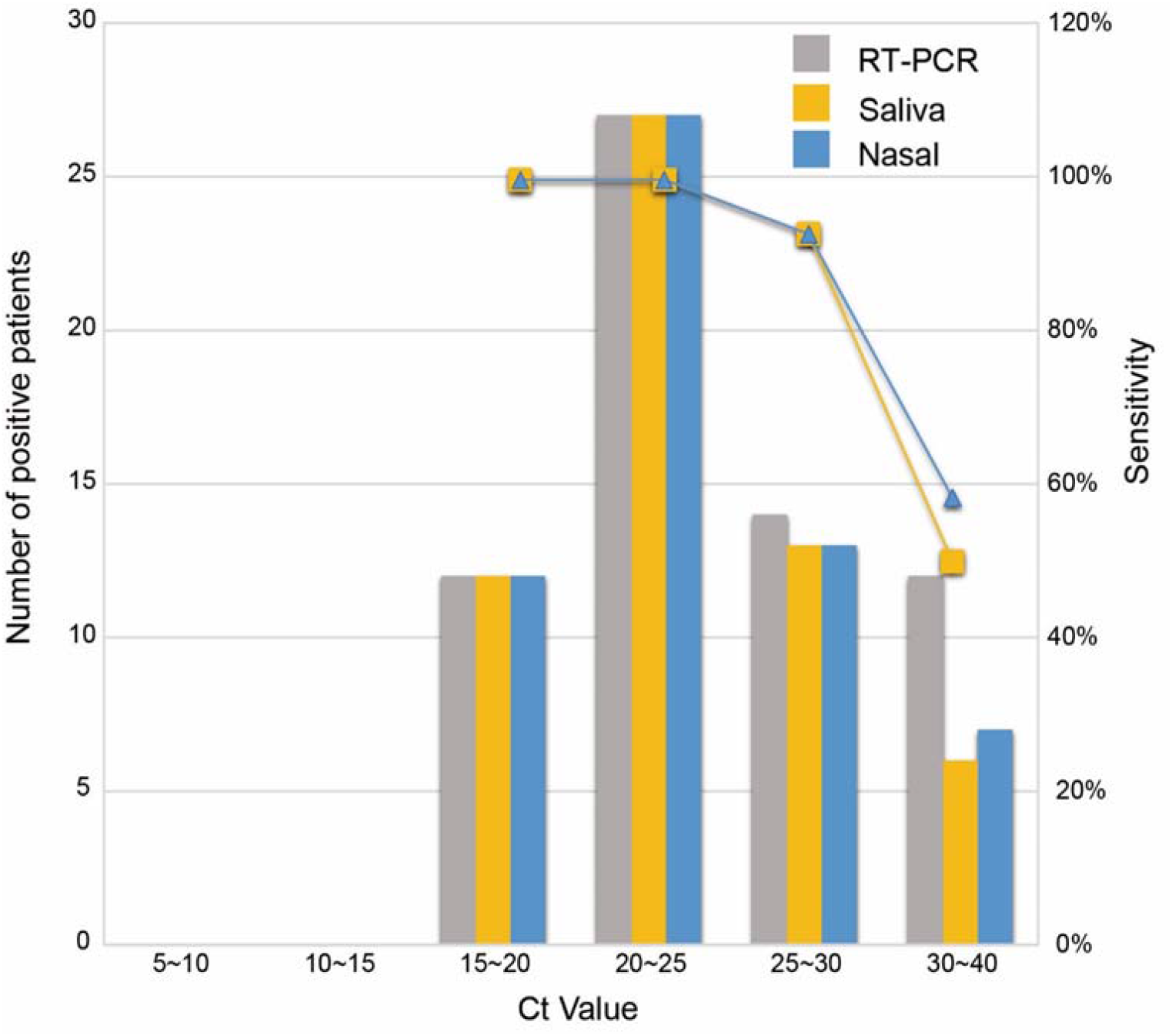
**Comparison of the sensitivity of PCL COVID19 Ag Gold (COV04S) depending on Ct values of RT-PCR in 65 COVID-19 positive patients, who provided both saliva and nasal specimens**.

## DISCUSSION

In this clinical study, we tested the performance of saliva and nasal sampling methods using PCL COVID19 Ag Gold self-test, and found that the saliva sampling method shows near identical results to the nasal swabbing method. The limitation of the saliva self-test is equal to that of the nasal self-test; as seen in Figure 2, the performance suffers when samples with low viral load are tested, which correlates to high Ct value, more number of days since symptom onset, and asymptomatic subjects. As seen in Figure 4, For samples with Ct values of 25 to 30, the sensitivity of the saliva sampling method decreases to around 90% and for samples with Ct values of 30 to 40, the sensitivity decreases to 50%. The performance of saliva antigen tests is highly dependent on viral load compared to RT-PCR, so it is best used for the purpose of screening symptomatic individuals early in the disease progression. For this reason, the manufacturer of the saliva tests clearly recommends the test be used 8 days within onset of symptoms. Clinical studies of this saliva sampling method on subjects after 8 days since symptom onset should be discouraged, as the manufacturer does not claim to diagnose subjects with low viral load with high sensitivity.

Nevertheless, the performances of the saliva test and the nasal test are comparable, suggesting that they may be interchangeable according to the preference of the testing subject. Saliva sampling is less invasive than nasopharyngeal or nasal sampling, making it more viable for mass screening of children under the age of 18. Our clinical study is, to our knowledge, first to include children between the ages of 2 to 18, showing the potential for use in frequent mass screening at schools as a practical and effective alternative to the costly RT-PCR or the more invasive nasal antigen tests. Even with the lower sensitivity for samples with low viral loads, the rapid antigen kits may be a useful first-line-of-defense when adopted to the wide population for frequent testing, considering the low cost compared to the RT-PCR; among asymptomatic subjects, the saliva self-test detected Covid-19 positive patients with sensitivity of around 80%.

In result, our study supports that the sensitivity and specificity of PCL COVID19 Ag Gold self-test using saliva specimen are commensurate with those of rapid antigen self-test using nasal swab specimen, as well as the gold standard of RT-PCR test using NPS. Because saliva sample collection is more comfortable and non-invasive compared to nasal and NPS, PCL COVID19 Ag Gold self-test using saliva is a preferred alternative to RT-PCR test using NPS for frequent self-testing towards a large population.

## MATERIALS AND METHODS

### Study design and participants

In this prospective clinical evaluation study, fresh human saliva, nasal, and nasopharyngeal specimens were collected from 338 individuals enrolled in the study at Mohammed VI University of Health Sciences (UM6SS) between June 30 and August 6 of 2021. A study overview design schematic can be found in figure 5. Saliva and nasal swab specimens were collected and analyzed with rapid antigen test kits by study participants themselves or assisted by an adult for age group under 14 (nasal swab specimen) and age group under 10 (saliva specimen). Nasopharyngeal swab specimens were collected by the healthcare professionals and analyzed with the RT-PCR as a comparator test. The study was approved under the Institutional Review Board of Mohammed VI University of Health Sciences (UM6SS) (IRB No. CERB/UM6SS/05/21) and informed consents were all obtained from study participants. The details of inclusion and exclusion criteria are explained in supplemental material 1.

**Figure 5.**
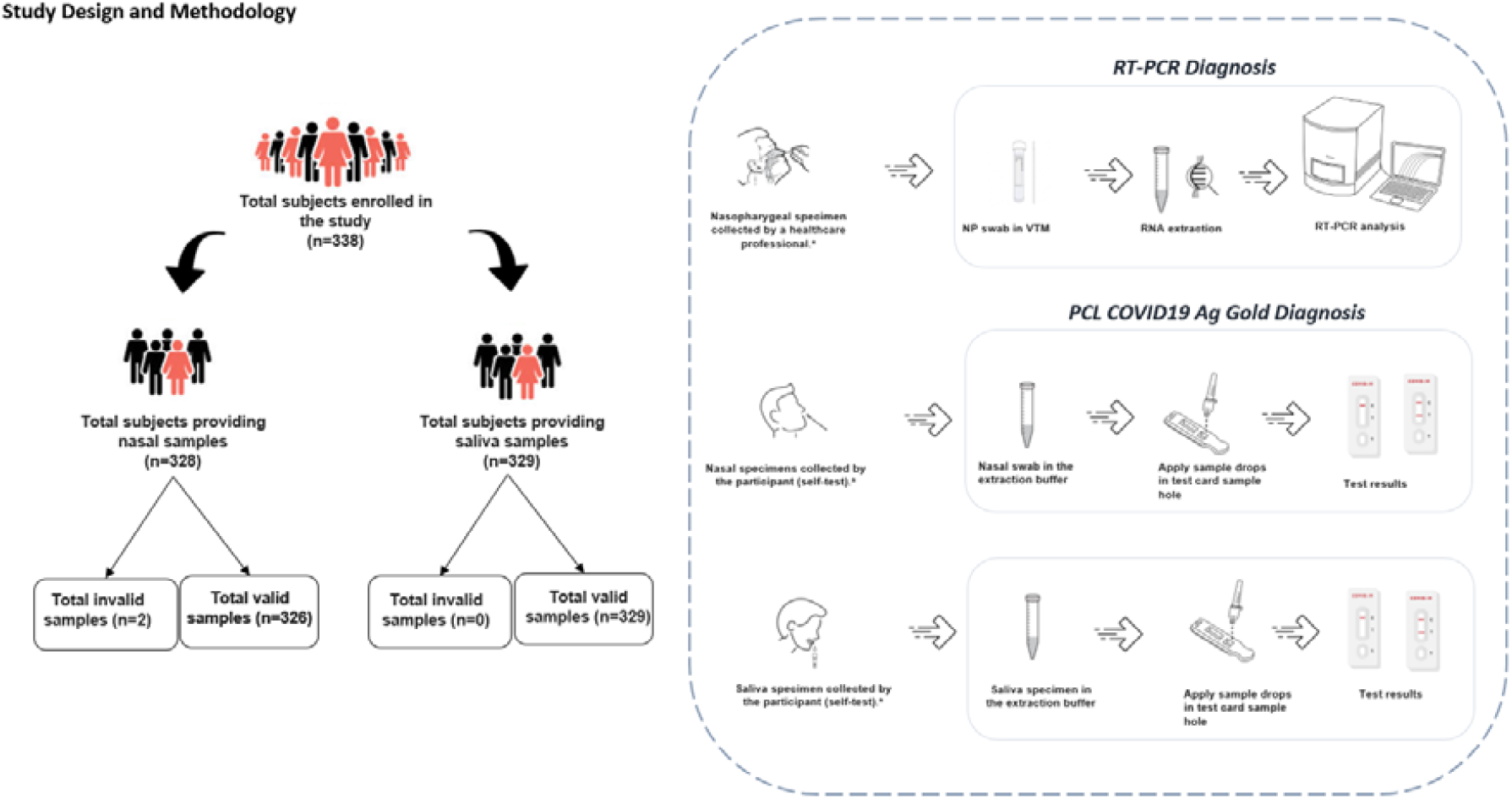
Clinical Study Workflow Schematics.

### Procedures

Eligible patients are enrolled to participate in the study. They are informed of possible risks of the procedure and are required to give informed consent before study-specific procedures can be done. To ensure the randomization and blinding of the tests, all enrolled subjects, or testers testing another individual, performed the entire study procedure in a private area that resembles a home setting without any assistance from the study personnel. The order of obtaining nasal swab and saliva specimens was randomized to ensure unbiased results. Participants were instructed not to eat or drink any beverages apart from water before the specimen collection. Either nasal swab specimen or saliva specimen or even both were tested depending on the subject’s preference and consent for the PCL COVID19 Ag Gold self-test following the product’s IFU. For nasal swab specimen, if the subject’s age was under 14 then the test was assisted by the adult and for saliva specimen, if the subject’s age was under 10 then the test was assisted by the adult as well. Nasopharyngeal swab specimen was also obtained as a comparator sample from the subject by the healthcare professional for the RT-PCR comparator test (US FDA Emergency Use Authorized). The entire rapid test process including sample collection, testing, and interpretation was performed by each individual study subject using the materials provided from the kit. The study subjects were observed by study personnel during the procedure and all difficulties were noted.

### Statistical analysis

The statistical analysis was done on R using ‘caret’ package.

## Data Availability

All data produced in the present study are available upon reasonable request to the authors.

